# An International, Cross-Sectional Survey of Preprinting Attitudes Among Biomedical Researchers

**DOI:** 10.1101/2023.09.17.23295682

**Authors:** Jeremy Y. Ng, Valerie Chow, Lucas J. Santoro, Anna Catharina Vieira Armond, Sanam Ebrahimzadeh Pirshahid, Kelly D. Cobey, David Moher

**Affiliations:** Centre for Journalology, Ottawa Methods Centre, Ottawa Hospital Research Institute, Ottawa, Ontario, Canada; Department of Health Research Methods, Evidence, and Impact, Faculty of Health Sciences, McMaster University, Hamilton, Ontario, Ontario, Canada; School of Epidemiology and Public Health, Faculty of Medicine, University of Ottawa, Ottawa, Canada; Meta-Research and Open Science Program, University of Ottawa Heart Institute, Ottawa, Canada

## Abstract

**Background:** Preprints are scientific manuscripts that are made available on open-access servers but are not yet peer reviewed. While preprints are becoming more prevalent uptake is not uniform or optimal. Understanding researchers’ opinions and attitudes towards preprints is valuable to their successful implementation. Understanding knowledge gaps and researchers’ attitudes toward preprinting can assist stakeholders like journals, funding agencies, and universities to implement preprints more effectively. Here, we aim to collect perceptions and behaviours regarding preprints in across an international sample of biomedical researchers.

**Methods:** Biomedical authors were identified by a keyword-based, systematic search from the MEDLINE database, and their emails were extracted to invite them to our survey. A cross-sectional anonymous survey was distributed to all identified biomedical authors to collect their knowledge, attitudes, and opinions about preprinting.

**Results:** The survey was completed by 730 biomedical researchers with a response rate of 3.20% and demonstrated a wide range of attitudes and opinions about preprints with authors from various disciplines and career stages around the world. Most respondents were familiar with the concept of preprints, but most had not published a preprint before. The lead author of the project and journal policy had the most impact on decisions to post a preprint, while employers/research institute had the least impact. Supporting open science practices was the highest ranked incentive, while increases to authors’ visibility was highest ranked motivation for publishing preprints.

**Conclusion:** While many biomedical researchers recognize the benefits of preprints, there is still hesitation among others to engage in this practice. This may be due to the general lack of peer review of preprints and little enthusiasm from external organizations, such as journals, funding agencies, and universities. Future work is needed to determine optimal ways to increase researcher’s attitudes through modifications to current preprint systems and policies.

## Background

The term “preprints” refers to scientific manuscripts that are made available on an open-access infrastructure, known as the “preprint server”, that have not yet undergone formal peer review^1,2^. One of the primary purposes of preprints is to make research available as quickly as possible, given the time lag from journal submission to publication, which typically takes around 8 to 10 months in the biomedical sciences due to multiple review rounds, editorial decision-making, high rejection rates, and time needed for revisions^2–6^. Preprints do not rely on peer review prior to release, allowing knowledge to be shared quickly with the introduction of novel results and methodology that can save months to years of research time^4^. Preprints have become incredibly valuable for authors as a larger group of researchers can critique their work (i.e., open peer review) with the preprint servers’ feedback system that allows for public and open feedback directly onto a manuscript that encourages discussion^7^. The FAST (Focused, Appropriate, Specific and Transparent) principles, which are guidelines that reviewers can follow when reviewing preprints, allow for high quality, constructive feedback to be provided^8^. As a result, researchers can improve their final manuscript, so it can possibly be published sooner and with fewer revisions^3^. In addition, authors have also begun to more frequently cite preprints in their working manuscripts as institutions and funders become more open and permissible in their policies regarding preprints^3^. Furthermore, funders acknowledge the importance of preprints by encouraging researchers to post and reference their preprints in their grant applications^3^. Preprint servers still utilize a minimal screening process to evaluate articles for incompleteness, plagiarism, and if they blatantly disregard or contradict widely accepted medical practices that could potentially jeopardize someone’s health if posted^9^.

However, many scientists are still hesitant about preprinting despite their benefits. Some major concerns include the credibility of preprints and premature media coverage^3,6,10^. With the COVID-19 pandemic, online media used preprints to introduce novel research to the public rapidly. However, patient and the public may be less familiar with the fact that preprints have not yet undergone the peer-review process and may fail to discern the difference between a preprint and an article published in the peer-reviewed literature^3,10^. Nonetheless, much COVID-19 research posted on preprint servers received high levels of coverage, and certain preprints could be used to push certain agendas, such as conspiracy theories and xenophobia^10^. This has led to the potential for the spread of misinformation^10,11^.

The purpose of this study is to explore biomedical researchers’ attitudes towards preprinting, through the administration of an anonymous cross-sectional online survey. We aim to determine what factors influence biomedical researchers’ opinions about publishing and viewing preprints. From this, future work may focus on determining how to improve researchers’ attitudes toward preprinting. In addition, our results may potentially impact how other stakeholders implement and modify their preprint policies in the future.

## Methods

### Approach

Prior to commencing this study, a protocol was registered on the Open Science Framework (OSF)^12^ before participant recruiting began^13^. The study used the MEDLINE database to identify participants for our survey using publicly available information. Once identified, participants were invited to complete an online survey. Survey questions were purpose-built for the survey by a core team of authors. They focused on understanding biomedical researchers’ attitudes and opinions concerning preprinting. This study was approved by the Ottawa Health Sciences Research Ethics Board (OHSN-REB Number: 20220584-01H).

### Open Science Statement

The study materials and raw data have been made available via OSF at the time of publication submission.

### Study Design

We conducted an anonymous, online, cross-sectional closed survey of published authors within biomedical journals.

### Sampling Framework

The sampling methods followed those described in Ebrahimzadeh et al.^14^. We utilized and downloaded the MEDLINE database of journals which contained approximately 30,000 journals. From this list, 400 journals were randomly selected using the RAND() function in Microsoft Excel. Authors’ names and emails were extracted using our script from articles published from 2021/07/01 to 2022/08/01. **Supplementary File 1** outlines the semi-automated process we used, generated by Ebrahimzadeh et al.^14^.

### Participant Recruitment

Only researchers who have been identified by our sampling framework developed by Ebrahimzadeh et al.^14^ received the closed survey. Using SurveyMonkey^15^, prospective participants received an email invitation to complete the survey. The email included an authorised recruitment script that described the study and its goals, invited recipients to review our informed consent form, and asked them to complete our anonymous online survey. Upon clicking the survey link in the invitation email, participants had to read and agree with the informed consent form before being able to see the survey. The initial list of 24 000 names and emails of corresponding authors contained duplicates and no longer functioning emails. We were able to determine no longer functioning emails due to SurveyMonkey’s bounced function, which labels certain contact information that have a permanent reason for an email or text to not be delivered. Before recruiting, duplicate authors and no longer functioning emails were deleted from the dataset. Therefore, we emailed 22 808 corresponding authors. We predicted that we would receive about 2200 responses based on a predicted response rate of around 10%. There was no financial compensation and no requirement to participate. Anyone who did not want to respond to a question could skip it.

### Survey

The complete survey is available in **Supplementary File 2**. The survey was created, pilot tested by SEP, DM, KC, and ACA, distributed, and collected using the University of Ottawa’s SurveyMonkey account^15^. The survey contained 29 questions and SurveyMonkey estimated that it took approximately 15 minutes to complete. The survey initially asked participants 5 general demographic questions including gender, research role, area of research, and country of residence. They were then asked 9 questions about their experiences with preprints, and 15 questions about their preferences and opinions of preprinting. The survey used adaptive formatting, which means that each participant’s prior response will influence the next question they see and the answers that might be provided. Most of the questions were multiple-choice; the remaining were open-ended, requiring participants to write their responses in a text box. Reminder emails were sent to participants after the first and second week following the original invitation. The survey was closed approximately five weeks after the initial invitation email was sent. Both the initial invitation and the reminder emails made this clear to the survey takers. Responses were collected from January to February 2023.

### Data Management and Analysis

In accordance with Ebrahimzadeh et al.^14^, the survey data that was gathered from the participants were exported to Excel. Basic descriptive statistics like count and percentages were generated based on the analysis of the quantitative data. Based on gender, employment sector, current career stage, and research area, the information and replies from participants were compared and studied. Thematic content analysis was used by different members of the research team to individually code the replies to the specific qualitative items. To reach an agreement on respective codes, which are categorically classified and specified into distinct tables, researchers carried out multiple rounds of discussion. The Checklist for Reporting Results of Internet E-Surveys (CHERRIES) was used to inform the reporting of this survey^16^.

## Results

### Demographics

Our survey received 730 responses, with a response rate of 3.20% and completion rate of 95.48%. Incomplete responses were defined as responses with no questions answered after the second page of the survey. Moreover, we have reported our raw response rate, which is underestimated as we cannot determine how many of the 22 808 authors who were emailed currently identify as a biomedical researcher or had an actively working email address. All questions were optional and therefore, we provide the total number of respondents for each question. The provided percentages were appropriately calculated using each questions’ total number of responses. Over three-fifths of respondents (n=455, 62.41%) identified as senior researchers, which we defined as researchers who started their careers after formal education over ten years ago. Of the 729 responses, participants identified as the following: academia (n=580, 79.56%), research staff with no formal academic or industry position (n=58, 7.96%), government scientist (n=28, 3.84%), third sector (n=8, 1.10%), pharmaceutical industry (n=5, 0.69%), and none of the participants was part of the scholarly communication industry. We received 50 responses (6.86%) that chose the “other” option, which mainly consisted of participants that were part of the biotechnology industry, retired personnel, clinical researchers, clinicians, medical practitioners, and students. Detailed demographics and other aggregate participant data are shown in **Table 1**. The responses from participants were also categorized based on gender, employment sector, current career stage, and research area, as found here:.

**Table 1:** Characteristics of Survey Respondents.

|  |  |
| --- | --- |
| Gender (n=728) |  |
| Male | 401 (55.08%) |
| Female | 312 (42.86%) |
| Prefer not to indicate | 14 (1.92%) |
| Non-binary | 1 (0.14%) |
| Country of Primary Affiliation (n=698) |  |
| United States of America | 189 (27.08%) |
| Canada | 69 (9.89%) |
| India | 41 (5.87%) |
| United Kingdom of Great Britain and Northern Ireland | 38 (5.44%) |
| Australia | 34 (4.87%) |
| Spain | 32 (4.58%) |
| China | 29 (4.15%) |
| Italy | 25 (3.58%) |
| Brazil | 18 (2.58%) |
| Netherlands | 18 (2.58%) |
| Iran (Islamic Republic of) | 16 (2.29%) |
| Turkey | 14 (2.01%) |
| Germany | 12 (1.72%) |
| Switzerland | 12 (1.72%) |
| France | 11 (1.58%) |
| Denmark | 9 (1.29%) |
| Greece | 9 (1.29%) |
| Mexico | 8 (1.15%) |
| Japan | 7 (1.00%) |
| Norway | 7 (1.00%) |
| Austria | 6 (0.86%) |
| Republic of Korea | 6 (0.86%) |
| Belgium | 5 (0.72%) |
| Colombia | 5 (0.72%) |
| Poland | 5 (0.72%) |
| Romania | 5 (0.72%) |
| Sweden | 5 (0.72%) |
| Indonesia | 4 (0.57%) |
| Portugal | 4 (0.57%) |
| Russian Federation | 4 (0.57%) |
| Thailand | 4 (0.57%) |
| Chile | 3 (0.43%) |
| Ireland | 3 (0.43%) |
| Pakistan | 3 (0.43%) |
| United Arab Emirates | 3 (0.43%) |
| Albania | 2 (0.29%) |
| Bangladesh | 2 (0.29%) |
| Czech Republic | 2 (0.29%) |
| Jordan | 2 (0.29%) |
| Malaysia | 2 (0.29%) |
| New Zealand | 2 (0.29%) |
| Other* | 17 (2.38%) |
| **Current Career Stage (n=729)** |  |
| Senior researcher | 455 (62.41%) |
| Mid-career researcher | 139 (19.07%) |
| Early career researcher | 89 (12.21%) |
| Other (please specify) | 25 (3.43%) |
| Graduate student | 21 (2.88%) |
| **Current Employment Sector (n=729)** |  |
| Academic | 580 (79.56%) |
| Research staff (no formal academic/industry position) | 58 (7.96%) |
| Other (please specify) | 50 (6.86%) |
| Government scientist | 28 (3.84%) |
| Third sector (E.g., NGO, non-profit) | 8 (1.10%) |
| Pharmaceutical industry | 5 (0.69%) |
| **Current Research Area (n=727)** |  |
| Clinical research | 335 (46.08%) |
| Other (please specify) | 125 (17.19%) |
| Pre-clinical research - in vivo | 77 (10.59%) |
| Health systems research | 74 (10.18%) |
| Pre-clinical research - in vitro | 72 (9.90%) |
| Methods research | 44 (6.05%) |
| *Total of countries with 1 response |  |

### Experience

#### Familiarity with Preprints

We asked how familiar the participants were with the concept of a preprint based on the given definition, “A publicly available version of any type of scientific manuscript/research output preceding formal publication” ^17^. Of the 694 responses, participants ranked how familiar they were with the concept of preprints as such: very familiar (n=167, 24.06%), familiar (n=269, 38.76%), and somewhat familiar (n=92, 13.26%). Only 61 (8.79%) respondents had never heard of the term.

#### Publishing Experience

We then asked about participants’ personal experiences with posting on preprint servers. Of the 694 responses, approximately one-third of them (n=237, 34.15%) had authored more than 31 publications in the past 5 years. Most participants had not posted a manuscript on a preprint server, with 540 (78.15%) participants of 691 responses not posting their most recent work on a preprint server, and 414 (59.74%) respondents of 693 responses having never posted a manuscript on a preprint server before. However, when asked if the participants would create a preprint in the future, we received 695 responses: 303 (43.60%) participants said they would not, 296 (42.59%) participants said they were unsure if they would, and 96 (13.81%) participants said they would create a preprint in the future.

Of the 335 respondents that had posted preprints before, the most common preprint server used is bioRxiv, with 148 (44.18%) respondents publishing previously on this server. In addition, we asked at what point in the publication process are preprints posted, and of the 294 respondents that have previously posted preprints, 110 (37.41%) participants submitted their manuscripts prior to submitting them to a journal, while 133 (45.24%) participants posted a preprint simultaneously while submitting to a journal.

#### Viewing/Downloading Preprints

Interestingly, despite most participants not publishing their own works on preprint servers, more than two-thirds of 695 respondents (n=481, 69.21%) have previously viewed or downloaded preprints, though only approximately a quarter (n=176, 25.32%) had cited a preprint. The most common preprint server used to view/download preprints amongst 682 responses was bioRxiv (n=243, 35.63%), MedRxiv (n=148, 21.70%), and ResearchGate (n=140, 20.53%).

#### Peer Reviewing Preprints

We asked if peer review should become a part of the preprinting process, and 684 responded, with 231 (33.77%) respondents believing that peer review should be part of the preprint process, while 240 (35.09%) respondents said it should not. On the other hand, 213 (31.14%) were unsure **(Table 2).** We also asked if our participants had experience with peer reviewing a preprint and from the 688 responses, the vast majority (n=638, 92.73%) stated they had not. Of those that had peer reviewed a preprint, we wanted to know if the FAST principles were used during the process, and we received 205 responses. 148 (72.20%) respondents stated they were not familiar with the FAST principles, 44 (21.46%) said they did not use the FAST principles, and 13 (6.34%) said they did.

**Table 2:** Respondent’s Opinion on Mandating Peer Review in the Preprinting Process (n=684)

|  |  |
| --- | --- |
| Yes | 231 (33.77%) |
| No | 240 (35.09%) |
| I am not sure | 213 (31.14%) |

We also asked if participants believed that patients or members of the public should be able to peer review preprints. Of the 445 who expressed an opinion, 320 (71.91%) believed that patients and members of the public should not be able to peer review preprints, while 125 (28.09%) believed they should be able to.

### Factors and Attitudes towards Preprinting

We then wanted to determine what factors impact an author’s decision to post a preprint. We asked participants to assess which had the most significant impact on posting a preprint: employer/research institution, funding agency, lead author, co-author consensus, or journal policy. From the 651 respondents, the lead author has the most significant impact on the decision of posting a preprint, followed by journal policy, co-author consensus, funding agency, and the least impactful is the employer/research institution **(Figure 1).**

**Figure 1:**
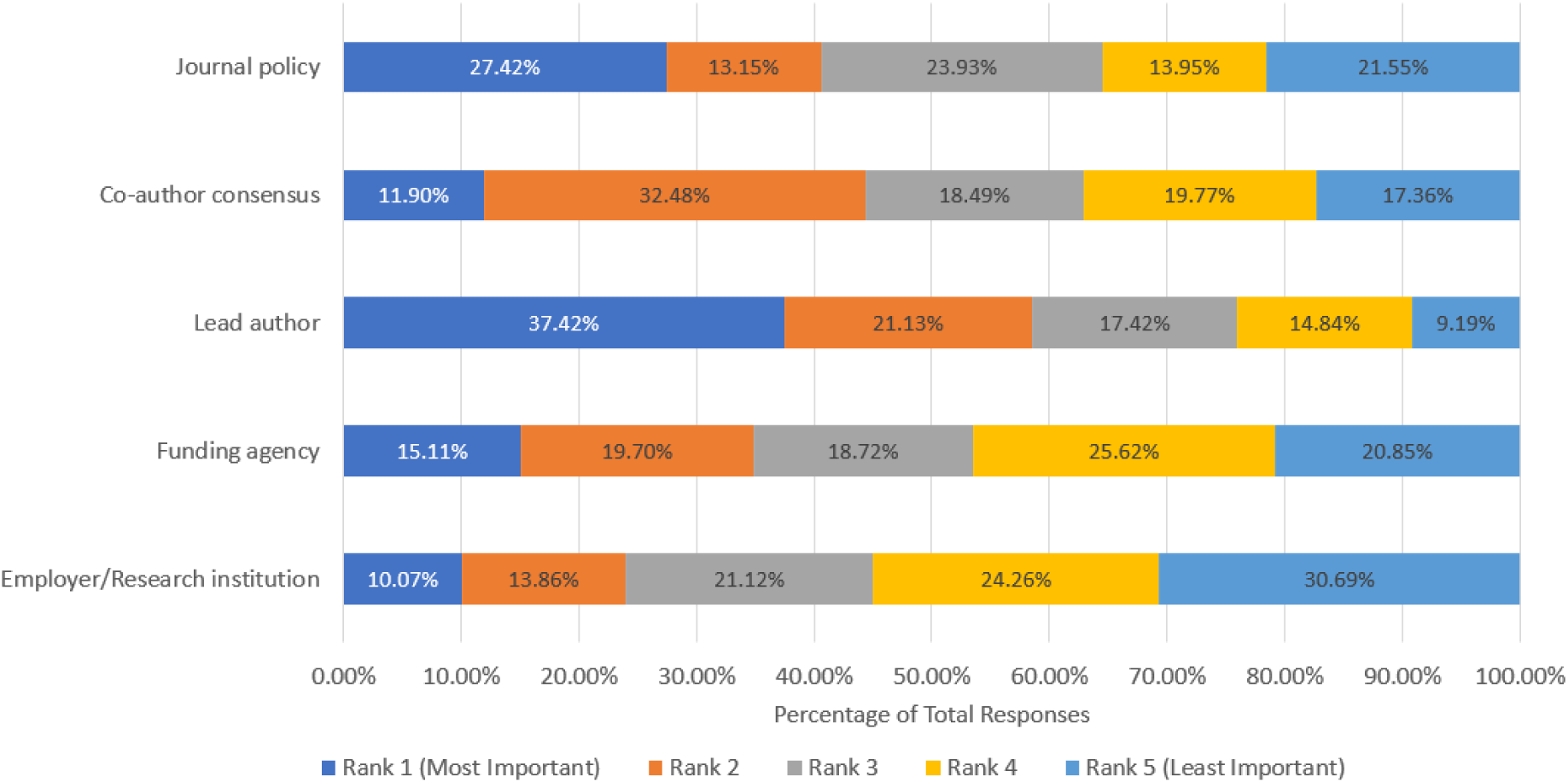
Ranking of Factors that Influence Respondents’ Decision to Post a Preprint.

We also asked if employers required or prohibited the posting of a preprint prior to submitting to a journal. Most employers do neither, with 649 (94.06%) respondents of 690 responses stating that their employer does not require them to post a preprint and 657 (95.08%) respondents of 691 responses stating that employers do not prohibit them from doing so. In addition, we wanted to see if a funder may impact this decision, and of 692 responses, 639 (92.34%) participants stated that funders do not require them to post a preprint.

We also wanted to determine how familiar participants were with the preprint policies of journals that they have experience publishing. First, we asked if participants were familiar with either Sherpa Romeo or Transpose Publishing, which are resources that present the preprint policies of most peer-reviewed journals. Of 686 responses, 623 (90.95%) respondents were not familiar with Sherpa Romeo and 656 (95.91%) respondents were not familiar with Transpose Publishing. In addition, we asked what the preprint policy was for the journals of the respondents’ most recent first author/co-author publication **(Figure 2).** From the 684 responses, 282 (41.23%) respondents stated that they were not familiar with the preprint policies, while 183 (26.75%) respondents stated that preprinting was permitted. Only 34 (4.97%) respondents and 61 (8.92%) respondents stated that preprinting was prohibited or had no preprinting policies, respectively.

**Figure 2:**
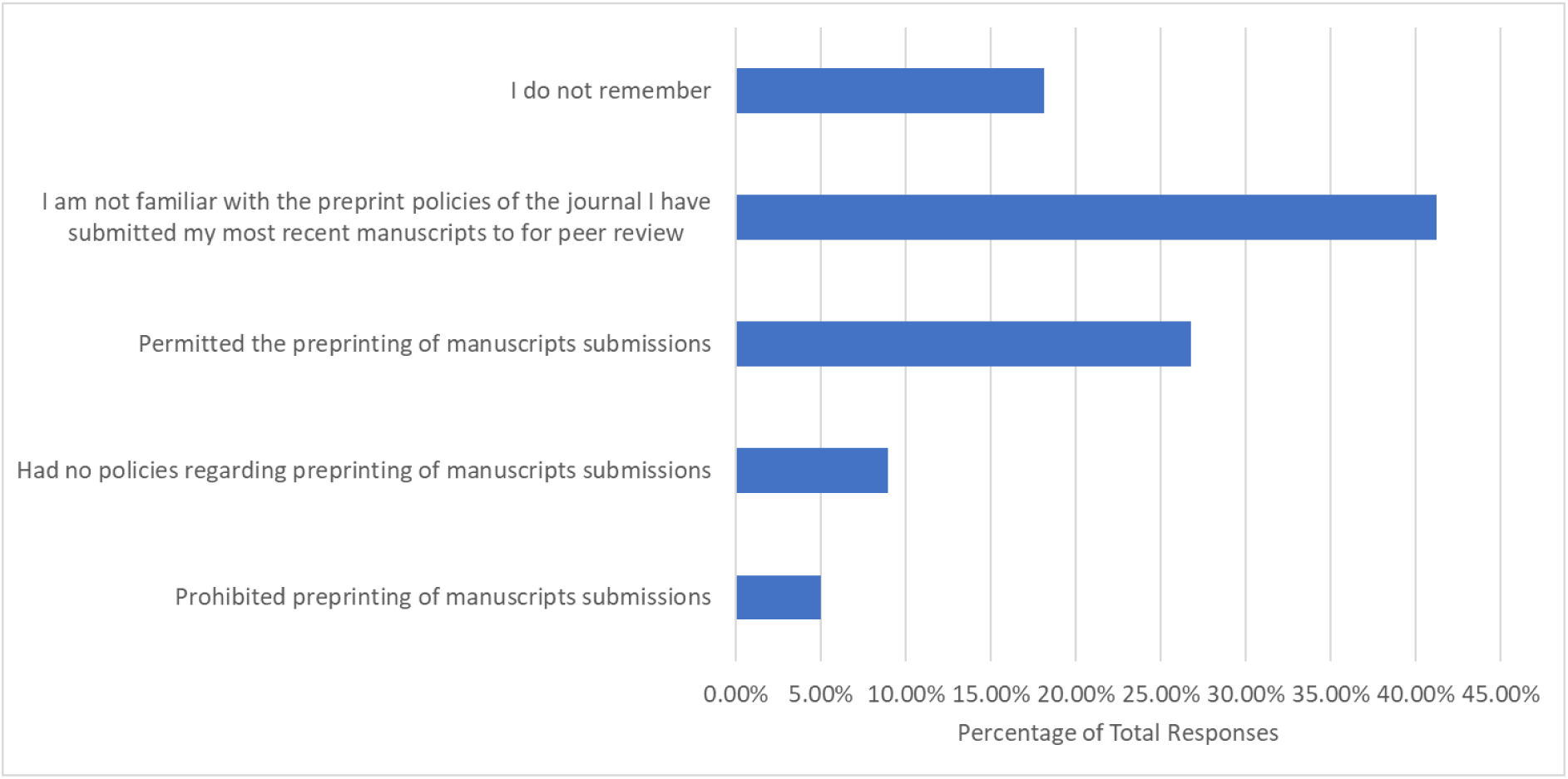
Preprint Policies of Journals Respondents’ had Most Recently Submitted a First Author/Corresponding Author Publication.

Respondents were asked to what extent they agreed with certain incentives or consequences of preprinting **(Figure 3).** Of the 666 who answered, over half had positive attitudes for the following incentives and consequences: 1) Preprints support open science practices (n=456, 68.99%), 2) Preprints should not be cited because they may contain results that lack credibility because they have not yet undergone peer review (n=433, 65.12%), 3) Preprints can be used as a weapon for the dissemination of misinformation (n=417, 62.61%), 4) Preprinting behaviors will increase in the biomedical field in the future (n=415, 62.32%), and 5) Preprinting allows for more efficient scientific dissemination (n=400, 60.06%). In terms of if respondents agreed or disagreed with the statement “Preprinting may encourage other researchers to steal project ideas (i.e., scooping), less than half (n=304, 45.66%) respondents agreed, while 143 (21.47%) respondents remained neutral, and 219 (32.89%) respondents disagreed. On the other hand, 284 respondents (42.71%) did not agree that preprinting would ultimately lead to higher-quality research being published, while 224 (33.68%) and 157 (23.60%) respondents chose neutral or agreement options, respectively.

**Figure 3:**
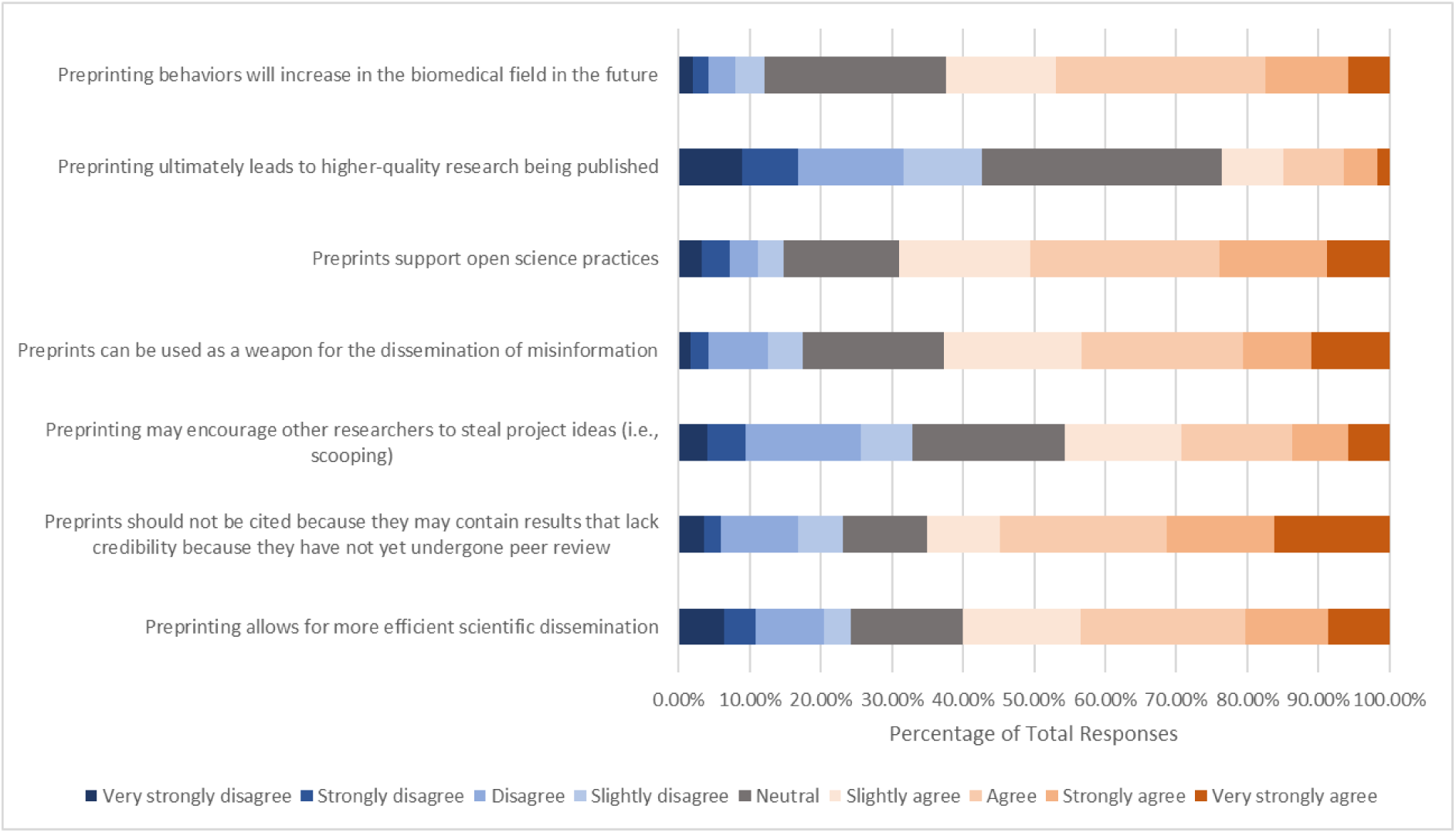
Respondents’ Opinions on Incentives and Consequences of Preprinting.

We then asked to what extent respondents agree with the following motivations to publish a preprint **(Figure 4).** Of the 666 respondents who answered, over half had positive attitudes toward the following incentives and consequences: 1) Preprinting increases authors’ visibility and allows for networking (n=489, 73.43%), 2) Preprinting allows for early/more feedback on their manuscript submission (n=470, 70.58%), 3) Preprinting allows authors to share studies that have been rigorously conducted but present negative/perceived low-impact results (i.e., lower likelihood of publication in a peer-reviewed journal) (n=437, 65.70%), 4) Preprinting makes my research visible to all if it is published in a paywall journal (n=415, 62.89%), and 5) Preprinting increases the likelihood of authors receiving more views and citations on their published article (n=365, 55.14%). In addition, slightly less than half of respondents (n=323, 48.57%) agreed that preprinting allows authors to prevent duplication of efforts, while 193 (29.02%) respondents disagreed, and 149 (22.42%) respondents remained neutral. In terms of if respondents agreed or disagreed with the statement “Preprints aid researchers’ careers with respect to hiring, promotion, and tenure”, respondents’ opinions were more evenly split, with 205 (30.91%) respondents agreeing, 218 (32.88%) respondents remaining neutral, and 240 (36.19%) respondents disagreeing.

**Figure 4:**
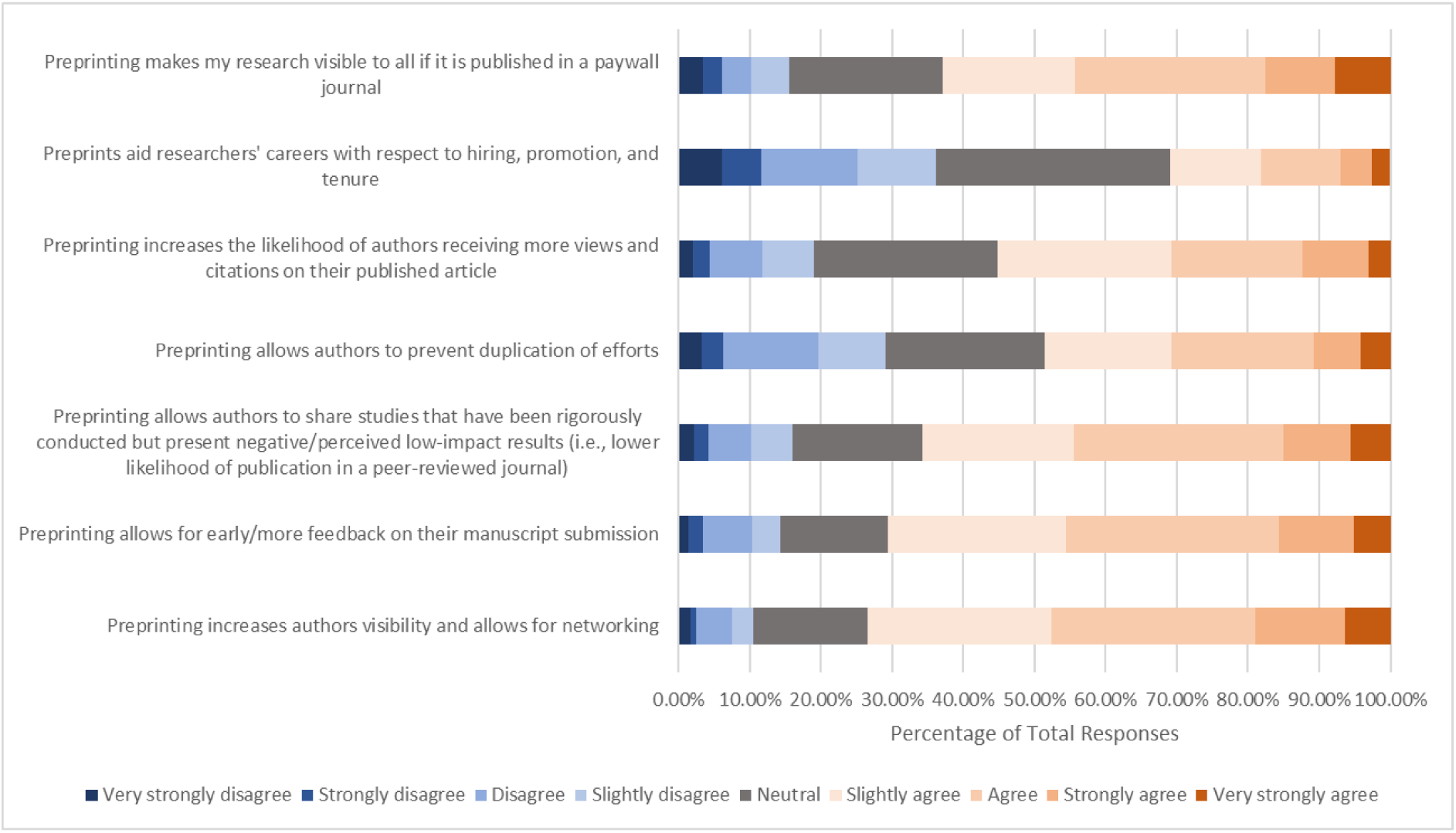
Respondents’ Opinions on Motivations to Publish a Preprint.

In addition, we asked participants for their opinion on making preprinting mandatory in the research process. Of the 665 respondents who expressed an opinion, over half (n=363, 54.58%) were leaning towards disagreeing with the statement. Only approximately a quarter (n=163, 24.52%) of respondents had positive opinions on this statement.

We then asked an open-ended question to allow respondents to freely express other factors that have motivated or dissuaded them from preprinting **(Table 3).** The 197 responses could primarily be split into positive factors and negative factors. The most prevalent positive factor that motivated preprinting “Preprints allow rapid dissemination”. This was followed by “Preprints show research efforts”, in which preprinting allowed authors to show their research to other parties, such as other researcher, funding agencies, and universities, without use of a publication in a journal. Lastly, the final positive factor commonly mentioned was “Preprints are open access items”.

**Table 3:**
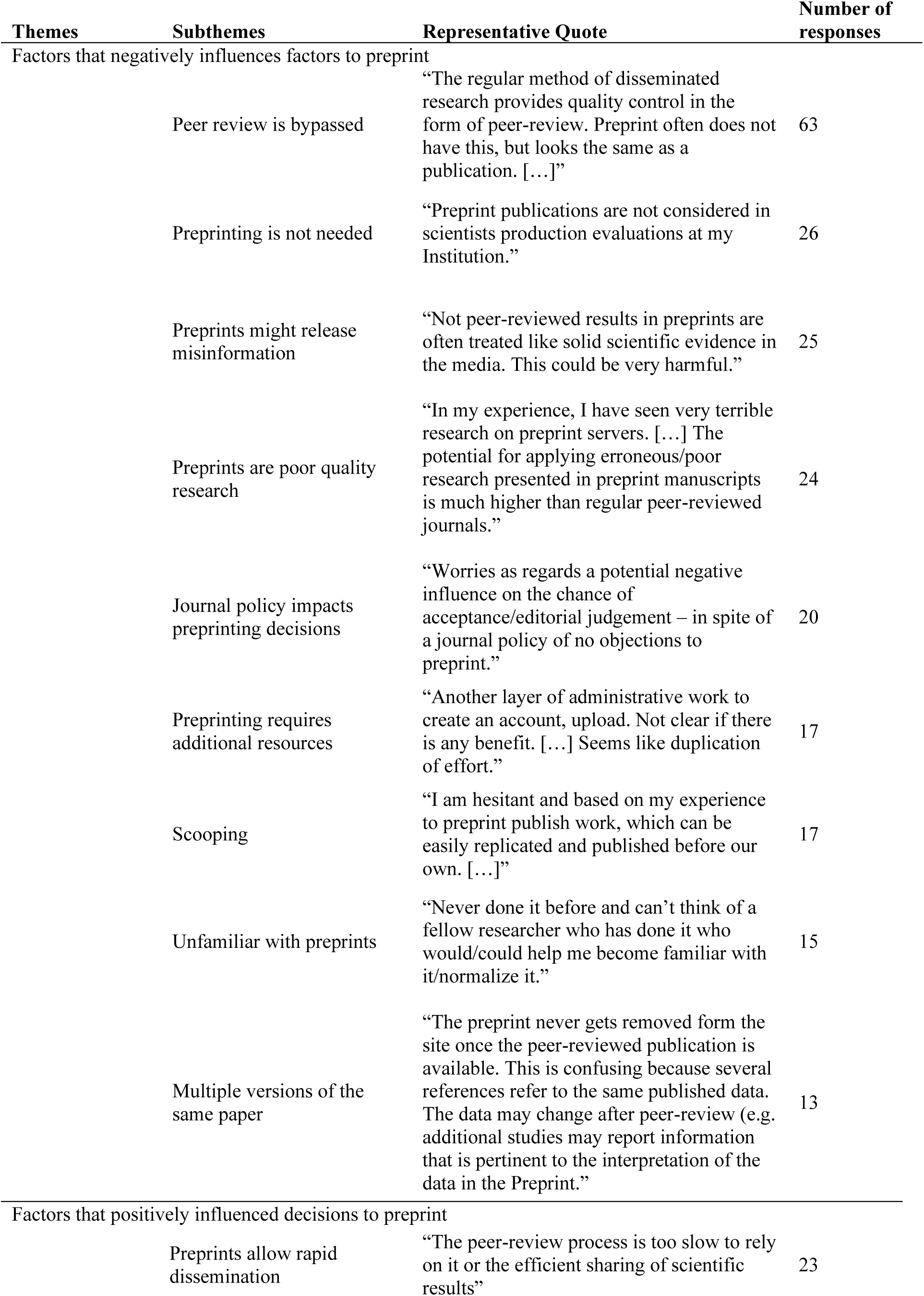
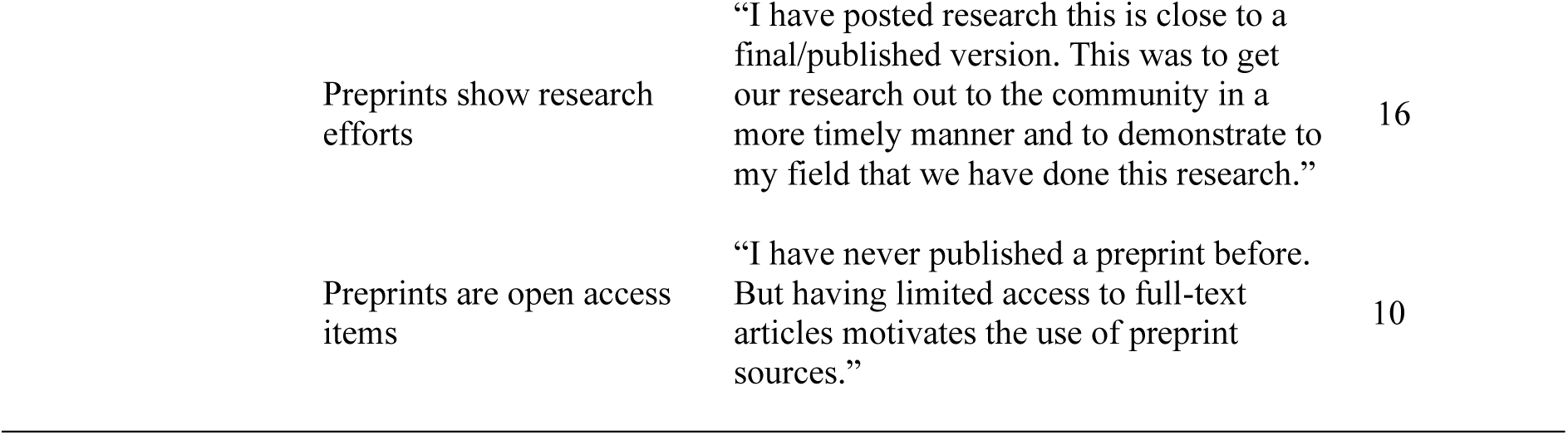
Thematic Analysis of the Open-ended Question (n=197)

| Themes | Subthemes | Representative Quote | Number of responses |
| --- | --- | --- | --- |
| Factors that negatively influences factors to preprint |  |  |  |
|  | Peer review is bypassed | “The regular method of disseminated research provides quality control in the form of peer-review. Preprint often does not have this, but looks the same as a publication. [...]” | 63 |
|  | Preprinting is not needed | “Preprint publications are not considered in scientists production evaluations at my Institution.” | 26 |
|  | Preprints might release misinformation | “Not peer-reviewed results in preprints are often treated like solid scientific evidence in the media. This could be very harmful.” | 25 |
|  | Preprints are poor quality research | “In my experience, I have seen very terrible research on preprint servers. [...] The potential for applying erroneous/poor research presented in preprint manuscripts is much higher than regular peer-reviewed journals.” | 24 |
|  | Journal policy impacts preprinting decisions | “Worries as regards a potential negative influence on the chance of acceptance/editorial judgement – in spite of a journal policy of no objections to preprint.” | 20 |
|  | Preprinting requires additional resources | “Another layer of administrative work to create an account, upload. Not clear if there is any benefit. [...] Seems like duplication of effort.” | 17 |
|  | Scooping | “I am hesitant and based on my experience to preprint publish work, which can be easily replicated and published before our own. [...]” | 17 |
|  | Unfamiliar with preprints | “Never done it before and can’t think of a fellow researcher who has done it who would/could help me become familiar with it/normalize it.” | 15 |
|  | Multiple versions of the same paper | “The preprint never gets removed form the site once the peer-reviewed publication is available. This is confusing because several references refer to the same published data. The data may change after peer-review (e.g. additional studies may report information that is pertinent to the interpretation of the data in the Preprint.” | 13 |
| Factors that positively influenced decisions to preprint |  |  |  |
|  | Preprints allow rapid dissemination | “The peer-review process is too slow to rely on it or the efficient sharing of scientific results” | 23 |

|  |  |  |
| --- | --- | --- |
| Preprints show research efforts | “I have posted research this is close to a final/published version. This was to get our research out to the community in a more timely manner and to demonstrate to my field that we have done this research.” | 16 |
| Preprints are open access items | “I have never published a preprint before. But having limited access to full-text articles motivates the use of preprint sources.” | 10 |

On the other hand, the open-ended questions showed that there were many negative factors that dissuaded respondents from preprinting. The most frequent, with approximately a third of responses, was “Peer review is bypassed”, as many responses state that peer review must be done to show credibility and quality of research before publishing. In addition, many responses believed that “Preprinting is not needed”, since preprinting does not provide any benefits to the individual’s career and there were no issues with the current publishing system. Another negative factor was “Preprint might release misinformation”, followed by “Preprints are poor-quality research”. Next, many responses stated that “Journal policy prevented preprinting”, since journal policy rejected or was unclear about preprints. “Scooping” was also mentioned frequently, along with “Preprinting requires additional resources”, where respondents felt that preprinting needed extra time, effort, and money that is unnecessary for overall publication of their work. Additionally, many respondents were “Unfamiliar with preprints” which discouraged them from preprinting. Lastly, “Multiple versions of the same paper” was another negative factor, where having a preprint and a journal published version of the same paper results in the release of unedited, possibly incorrect, work and causes confusion when referencing.

## Discussion

The objective of this study is to explore biomedical researcher’s attitudes towards preprint to determine what factors influence their opinions about publishing and viewing preprints. We found that the most impactful preventative factors against preprinting is the fear of disseminating misinformation, the lack of peer review, and unsupportive journal policies. While hiring, promotion, and tenure does not encourage preprinting, respondent do believe that preprints are beneficial in increasing visibility and recognition, being openly accessible, providing rapid feedback, and publishing negative data.

From this, future work may focus on determining how to improve researchers’ attitudes toward preprinting, and how other stakeholders can implement and modify their preprint policies in the future. This cross-sectional survey is, to our knowledge, the first that focused exclusively on biomedical researchers to explore their attitudes towards preprinting. The results of this approach are essential for gaining a clearer grasp on the perspectives that biomedical researchers have on preprints, which can potentially impact how other researchers, institutions, and publishing houses implement preprint policy in the future to improve preprinting in biomedical researchers.

### Opinions on the Benefits and Consequences of Preprints

In terms of benefits and consequences of preprints, the three statements that respondents most agreed with were: 1) Preprints support open science practices, 2) Preprints should not be cited because they may contain results that lack credibility because they have not yet undergone peer review, and 3) Preprints can be used as a weapon for the dissemination of misinformation. These results align with preliminary work done by Funk et al.^18^, which found that the most concerning issues with preprints from the perspective of respondents were premature media coverage and public sharing of information prior to peer review, while respondents believed that the top benefit of preprints is open access.

On the other hand, the top three most impactful factors among respondents with respect to deciding to publish a preprint included: 1) increasing author visibility and networking opportunities, 2) allowing for early and more feedback on manuscripts, and 3) publishing of negative or perceived low-impact results. The findings by Funk et al.^18^ somewhat align with our results as they found that only over half of the respondents found early feedback as “very beneficial” and were only seen as less impactful in comparison to open access and speed of dissemination. However, publishing of negative or perceived low-impact results was the second least impactful factor, only being seen as “very beneficial” by approximately one-third of respondents^18^. This variation may be due to more awareness of the bias journals have towards positive results, and why this is an issue for the scientific community. Nowadays, authors aim to only publish positive “groundbreaking” results, and journals often reject papers with negative or non-significant results to increase readers, citations, and submissions^19,20^. However, negative and non-significant results are vital to the scientific process, as they allow for collective self-correcting progress and ensure that resources are not wasted on the replication of failed research^20,21^. A survey by Echevarría et al.^22^ showed that majority of authors believe this information should be shared, though only a handful have ever tried to publish negative results. Preprints may be an appropriate alternative for publishing negative data as they can quickly share this information without the long peer review process, there is no chance of the manuscript being rejected, and there are no concerns with journal metrics. Future work may focus on gaining a better understanding of researchers’ attitudes toward publishing negative results and preprint practices, to ensure that negative results are appropriately disseminated within the scientific community.

One hesitation towards the use of preprints revolves around the fact that preprints have not undergone peer review, and therefore, the scientific content has not been validated before being released to the public. This is seen in our findings, in which almost two-thirds of our respondents believe that preprints should not be cited because results may lack credibility since there was no peer review. Also, over two-thirds of our respondents had viewed or downloaded a preprint, but only a quarter had ever cited one, implying the distrust in preprint content. Additionally, peer review being bypassed was the most frequent factor that negatively impacted respondent’s motivations to preprint, as seen in the open-ended question, alongside fear that preprints will disseminate misinformation and poor-quality research. Thus, we consistently observe that peer review is regarded as highly important and necessary in publishing science by respondents. Respondents’ hesitation towards preprints due to the lack of peer review and fear of misinformation indicate a lack of knowledge about the evidence regarding the quality of the published literature and the effectiveness of peer review. Work done by Zeraatkar et al.^23^ has shown that there was no evidence that preprints provided results that were inconsistent with peer-reviewed publications, specifically seen with COVID-19 treatments. MEDLINE is also currently indexing preprints, supporting research that has been posted to eligible preprint servers to be easily discovered and preserved^24^. In addition, it is important to note that peer review is not always effective due to inconsistency and bias, as many peer reviewers have not had standard training and many training opportunities and courses are not openly accessible online, inhibiting researchers from completing training on peer review^23,25–28^. Thus, this blind belief in peer review may negatively impact researcher’s opinions towards preprints.

Interestingly, despite many respondents distrusting preprints due to lack of peer review, many were divided on if peer review should become part of the preprint process. This may be because preprints allow for the rapid dissemination of research as there is a delay from journal submission to publication when undergoing peer review^2,6^. If peer review were to be incorporated into preprinting in the same way it is done for journals, then the work of peer reviewers would increase, and the delay would become much longer. Thus, preprint servers should be modified to improve the credibility of preprints, as suggested by Soderberg et al.^6^. The current preprint feedback system allows for public feedback and discussion, which may aid in credibility or results. However, authors often fear unfair criticism from competitors, harm to their reputation, or softened criticism due to the public nature of feedback^7^. Fortunately, the FAST principles can alleviate these issues^8^. From our findings, majority of respondents had not peer reviewed a preprint before, and those that had, most were not familiar with the FAST principles. Therefore, efforts must be made to educate researchers about these principles to promote feedback of preprints. Future work may aim to better understand researchers’ opinions on peer reviewing preprints and potentially provide other novel solutions, such that the benefits of peer review can be implemented to the preprinting process without the current fears of preprint feedback.

### External Factors that Play a Role on Opinions and Attitudes Towards Preprints

Journal policy is the second most impactful factor on the decision for authors to post a preprint and was frequently mentioned in the open-ended question. In addition, most respondents were not familiar with a journal’s preprint policy. Nowadays, most, but not all, life science journals accept preprinted manuscripts for submission, and some journals either do not have a preprint policy or have contradicting statements^4^. Biomedical journals should create clear and concise preprint policies that allow researchers to understand if they can publish preprints prior to or during submission to a journal. In addition, our data showed that most researchers do not know of resources on preprint policies, such as Sherpa Romeo and Transpose Publishing, which gathers journal policies on open access and preprinting. In the future, it would be best to educate researchers on these types of resources to address concerns that preprints negatively impact chances of publishing in a peer reviewed journal.

Respondents did not have strong opinions on if preprints aid researchers’ careers with respect to hiring, promotion, and tenure. In addition, our findings showed that universities and research institutes had the least impact on one’s decision to publish a preprint, and the majority of our respondents’ employers neither prohibited nor permitted posting preprints. This opinion may be because few universities consider preprints when hiring and for promotion^29^. Therefore, hiring, promotion, and tenure policies should be modified, as current methods are heavily reliant on quantitative metrics, which has been recognized as a flawed system^30^. Preprints may prevent hiring committee’s bias towards or against a journal, due to a journals’ impact factor or reputation^31,32^. Inclusion of preprints to hiring, promotion, and tenure policies may improve biomedical researchers’ attitudes towards preprinting.

### Strengths and Limitations

By implementing a cross-sectional survey for this project, we took a snapshot of our population of interest without having to follow them through time. This study was able to generalize among biomedical researchers through randomly surveying a large sample of biomedical researchers, who had varying opinions regarding preprint postings. A limitation of our study design was that it was written in English, so researchers without a working knowledge of English could not take the survey. In addition, as the present study was based on an English-speaking, international sample, so it does not consider the impact of national policies on attitudes and opinions toward preprinting. Another limitation with our sampling strategy is that the list of email addresses used potentially included inactive or invalid addresses, which may be a result of changing professions, retiring, or passing away. Furthermore, we did not consider autoreplies. Therefore, our response rate is underestimated. Our questionnaire is built on self-declared attitudes and preprint practices, which may not accurately capture independently confirmed practices. Inherent to the cross-sectional survey design, a final limitation also includes a low response rate, recall bias, in which participants do not correctly remember past events, and non-response bias, in which participants do not want to or cannot complete a survey question. Also, responses mostly reflect the opinions of senior academics, and there is little feedback from early career researchers. Therefore, another limitation is that early career researchers have different views and experiences.

## Conclusions

The present study created an online survey and determined biomedical researchers’ knowledge, experiences, and attitudes towards preprinting. We observed that biomedical researchers were familiar with the concept of preprints but lacked experience working with preprints. The various attitudes and opinions of biomedical researchers provided represent a valuable contribution to the field of academic publishing and the changes needed in the scientific community. To our knowledge, this is the first study of its kind that observes the attitudes and opinions of preprints in a discipline-specific manner. This study can therefore be used as a model for any other academic researchers from alternative disciplines that may have an interest in preprinting. It will be useful to determine attitudes and opinions in other disciplines to find methods to improve preprinting and researcher’s attitudes towards it. It is necessary to recognize the attitudes and opinions of biomedical researchers to provide suggestions to stakeholders to implement and improve preprinting.

## Data Availability

All data produced are available online on the Open Science Framework.

https://doi.org/10.17605/OSF.IO/WN92Q

## List of Abbreviations

FAST: Focused, Appropriate, Specific and Transparent
CHERRIES: Checklist for Reporting Results of Internet E-Surveys
OSF: Open Science Framework

## Declarations

### Ethics Approval and Consent to Participate

This study was approved by the Ottawa Health Sciences Research Ethics Board (OHSN-REB Number: 20220584-01H).

### Consent for Publication

All authors consent to this manuscript’s publication.

### Availability of Data and Materials

All relevant data and materials are included in this manuscript and on Open Science Framework. Available at: https://doi.org/10.17605/OSF.IO/WN92Q

### Competing Interests

The authors declare that they have no competing interests.

### Funding

This study was unfunded.

## Authors’ Contributions

JYN: designed and conceptualized the study, collected and analysed data, co-drafted the manuscript, and gave final approval of the version to be published.

VC: assisted with the collection and analysis of data, co-drafted the manuscript, made critical revisions to the manuscript, and gave final approval of the version to be published.

LJS: assisted with the collection and analysis of data, made critical revisions to the manuscript, and gave final approval of the version to be published.

ACVA: collection and analysis of data, made critical revisions to the manuscript, and gave final approval of the version to be published.

SEP: collection and analysis of data, made critical revisions to the manuscript, and gave final approval of the version to be published.

KDC: assisted with the design, collection and analysis of data, made critical revisions to the manuscript, and gave final approval of the version to be published.

DM: assisted with the design, collection and analysis of data, made critical revisions to the manuscript, and gave final approval of the version to be published.

## Supplementary Files

### Supplementary File 1: Search Strategy Extracted from Cobey et al. (2022)

We obtained a list of all journals indexed in MEDLINE together with their NLM ID. A random list of 1,000 journals was generated in Excel using the RAND() function.

#### Two strategies used to retrieve the articles

1. We searched for all articles published in each journal using the search strategy *1234567.jc.* where “1234567” is the NLM ID of the journal. Where NLM ID is not available, we will use the syntax “Name of journal”.nj. We ran searches for each journal separately. After each search, we sorted the results by Entry date (descending) and export the first 20 results.
2. We carried out a combined search for all journals using the following search strategy:

1. 1234567.jc.
2. 2345678.jc.
3. 3456789.jc.
4. or/1-3
5. limit 4 to dt=yyyymmdd-yyyymmdd

#### Three strategies used to obtain the email addresses of authors

1. All retrieved articles will be re-imported to EndNote/Zotero/Mendeley to retrieve PMID numbers. The list of PMID numbers was exported as an .csv file and inputted into an R script (built based on the easyPubMed package) to retrieve the authors’ name, affiliation institutions and email addresses.
2. In addition, we used the Find Full Text function in EndNote to retrieve PDF files of these articles, and ran these files in another R script for text recognition to extract email addresses.
3. Any articles where email addresses cannot be retrieved from both methods were manually screened.

Results from all three methods were combined into the final list and counter-checked by another author for potential errors before survey distribution.

### Supplementary File 2: Survey

#### Demographics

1. What career stage best describes you?

a. Graduate student
b. Early career researcher (<5 years of formally starting your career post formal education)
c. Mid-career researcher (5-10 years of starting your career post formal education)
d. Senior researcher (>10 years of starting your career post formal education)
2. What best describes your current position?

a. Academic
b. Research staff (no formal academic/industry position)
c. Pharmaceutical industry
d. Scholarly Communication industry (journals/publishing)
e. Third sector (E.g., NGO, non-profit)
f. Government scientist
g. Other, please specify
3. What is your gender?

a. Male
b. Female
c. Non-binary
d. Other
e. Prefer not to say
4. What country are you employed in? (Drop-down menu)
5. Which of the following describes your research area?

a. Clinical research
b. Pre-clinical research - *in vivo*
c. Pre-clinical research - *in vitro*
d. Health systems research
e. Methods research
f. Other, please specify

#### Preprinting Behaviours, Motivations and Attitudes

6. Are you familiar with the concept of preprint, defined as “A publicly available version of any type of scientific manuscript/research output preceding formal publication” by <u>FORRT</u>?

a. Not familiar
b. Somewhat familiar
c. Familiar
d. Very familiar
7. Have you ever posted a manuscript you are first author on on a preprint server?

a. Yes
b. No
c. Don’t remember
8. For your most recent manuscript publication that you are first author on, did you post the manuscript on a preprint server?

a. Yes
b. No
c. Don’t remember
9. In the future, I will create a preprint for all first author papers I seek to publish

a. True
b. False
c. Unsure
10. How many publications have you been the author of in the past 5 years?

a. 1-5
b. 6-10
c. 11-15
d. 16-20
e. 21+
11. Have you ever viewed/downloaded a preprint in which you were not an author?

a. Yes
b. No
c. Don’t remember
12. Have you ever cited a preprint work in which you were not an author?

a. Yes
b. No
c. Don’t remember
13. What preprint servers have you used to publish manuscripts in the past? (Drop-down menu)

a. Chosen from list (https://asapbio.org/preprint-servers)
b. Don’t remember
c. Other, please specify.
14. What preprint servers have you used to view manuscripts in the past? (Drop-down menu)

a. Chosen from list (https://asapbio.org/preprint-servers)
b. Don’t remember
c. Other, please specify.
15. At what point in the publication process do you typically post preprints?

a. Prior to submitting your manuscript to a journal
b. Simultaneously when submitting your manuscript to a journal
c. After submitting your manuscript to a journal
d. After receiving an initial decision from the journal
e. After your manuscript is accepted for publication in a journal but before it is published
16. Does your employer require you to post a preprint of your manuscript prior to it being published in a peer-reviewed journal?

a. Yes
b. No
c. Don’t know
17. Has your employer ever prohibited you from posting a preprint?

a. Yes
b. No
18. Has a funder of your research ever required you to post a preprint?

a. Yes
b. No
c. Sometimes
d. Don’t remember
19. Of the following, who has the greatest impact on whether a preprint is posted or not? Please order the following items, with (1) being the most impactful, and (5) being the least impactful.

a. Employer/research institution
b. Funding agency
c. Lead author
d. Co-author consensus
e. Journal policy
20. To what extent are you familiar with the following websites, which presents the preprinting policies of most peer-reviewed journals (1-not familiar, 2-somewhat familiar, 3-familiar, 4-very familiar)

a. Sherpa Romeo
b. Transpose Publishing
21. What was the preprint policy of the journal of your most recent first author/corresponding author publication?

a. Prohibited preprinting of manuscripts submissions
b. Had no policies regarding preprinting of manuscripts submissions
c. Permitted the preprinting of manuscripts submissions
d. I am not familiar with the preprint policies of the journal I have submitted my most recent manuscripts to for peer review
22. Peer review is not typically part of the preprint process. Should it be?

a. Yes
b. No
c. I am not sure
23. Have you ever peer reviewed a preprint?

a. Yes
b. No
24. If yes, have you used the FAST principles to help you?

a. Yes
b. No
c. I am not familiar with the FAST principles
d. I answered NO to the previous question
25. Do you think patients/members of the public should be able to peer review preprints?

a. Yes
b. No
c. I am not sure

Questions 26-29 have been categorized based off Table 1 in Atkins et al. (2017) doi: 10.1186/s13012-017-0605-9.

26. Reinforcement: To what extent do you agree with these incentives/consequences of preprinting? (1-very strongly disagree, 2-strongly disagree, 3-disagree, 4-slightly disagree, 5-neutral, 6-slightly agree, 7-agree, 8-strongly agree, 9-very strongly agree)

a. Preprinting allows for more efficient scientific dissemination
b. Preprints should not be cited because they may contain results that lack credibility because they have not yet undergone peer review
c. Preprinting may encourage other researchers to steal project ideas (i.e., scooping)
d. Preprints can be used as a weapon for the dissemination of misinformation
e. Preprints support open science practices
f. Preprinting ultimately leads to higher-quality research being published
g. Preprinting behaviors will increase in the biomedical field in the future
27. Motivation and Goals: To what extent do you agree with the following motivations to publish a preprint? (1-very strongly disagree, 2-strongly disagree, 3-disagree, 4-slightly disagree, 5-neutral, 6-slightly agree, 7-agree, 8-strongly agree, 9-very strongly agree)

a. Preprinting increases authors visibility and allows for networking
b. Preprinting allows for early/more feedback on their manuscript submission
c. Preprinting allows authors to share studies that have been rigorously conducted but present negative/perceived low-impact results (i.e., lower likelihood of publication in a peer-reviewed journal)
d. Preprinting allows authors to prevent duplication of efforts
e. Preprinting increases the likelihood of authors receiving more views and citations on their published article
f. Preprints aid researchers’ careers with respect to hiring, promotion, and tenure
28. To what extent do you agree that preprinting should be made mandatory in the research process in the future?

a. Very strongly disagree
b. Strongly disagree
c. Disagree
d. Slightly disagree
e. Neutral
f. Slightly agree
g. Agree
h. Strongly agree
i. Very strongly agree
29. Please describe any other factors that may have motivated or dissuaded you to post a preprint.

(Text box)

## Notes

### Competing Interest Statement

The authors have declared no competing interest.

### Clinical Protocols

https://doi.org/10.17605/OSF.IO/QA9GN

### Funding Statement

This study did not receive any funding.

